# Polygenic Risk Prediction using Gradient Boosted Trees Captures Non-Linear Genetic Effects and Allele Interactions in Complex Phenotypes

**DOI:** 10.1101/2021.07.09.21260288

**Authors:** Michael Elgart, Genevieve Lyons, Santiago Romero-Brufau, Nuzulul Kurniansyah, Jennifer A. Brody, Xiuqing Guo, Henry J Lin, Laura Raffield, Yan Gao, Han Chen, Paul de Vries, Donald M. Lloyd-Jones, Leslie A Lange, Gina M Peloso, Myriam Fornage, Jerome I Rotter, Stephen S Rich, Alanna C Morrison, Bruce M Psaty, Daniel Levy, Susan Redline, the NHLBI’s Trans-Omics in Precision Medicine (TOPMed) Consortium, Tamar Sofer

## Abstract

Polygenic risk scores (PRS) are commonly used to quantify the inherited susceptibility for a given trait. However, the standard PRS fail to account for non-linear and interaction effects between single nucleotide polymorphisms (SNPs). Machine learning algorithms can be used to account for such non-linearities and interactions. We trained and validated polygenic prediction models for five complex phenotypes in a multi-ancestry population: total cholesterol, triglycerides, systolic blood pressure, sleep duration, and height. We used an ensemble method of LASSO for feature selection and gradient boosted trees (XGBoost) for non-linearities and interaction effects. In an independent test set, we found that combining a standard PRS as a feature in the XGBoost model increases the percentage variance explained (PVE) of the prediction model compared to the standard PRS by 25% for sleep duration, 26% for height, 44% for systolic blood pressure, 64% for triglycerides, and 85% for total cholesterol. Machine learning models trained in specific racial/ethnic groups performed similarly in multi-ancestry trained models, despite smaller sample sizes. The predictions of the machine learning models were superior to the standard PRS in each of the racial/ethnic groups in our study. However, among Blacks the PVE was substantially lower than for other groups. For example, the PVE for total cholesterol was 8.1%, 12.9%, and 17.4% for Blacks, Whites, and Hispanics/Latinos, respectively. This work demonstrates an effective method to account for non-linearities and interaction effects in genetics-based prediction models.

## Introduction

In the last few years, genetics-based trait prediction using polygenic risk scores (PRS) have become increasingly popular. PRS are calculated as weighted sums of allele counts for variants that are associated with an outcome of interest and are used to quantify the inherited susceptibility for a given trait or disease (1). Traditionally, genome wide association studies (GWAS) are used to identify the univariate relationships between single nucleotide polymorphisms (SNPs) and a given phenotype. These univariate relationships are then used to construct the PRS (2).

The prediction models that use PRS are generally able to explain only a small percentage of the observed variance for a given trait (3), which could be due to several factors. Because they rely on univariate effect sizes derived from linear GWAS models, standard PRS as defined above do not account for potential non-linearities in the association between the genetic data and the outcome of interest. Additionally, additive PRS models do not account for interactions between SNPs, which are known to exist (4). One common strategy employed during the SNP selection stage of PRS construction is clumping, to exclude SNPs within a predefined distance of one another and levels of linkage disequilibrium (LD). Potential interactions are not usually taken into account by this approach – as in haplotypes (5) or epistatic effects (6) both inside and outside the clumping region. Examples of strong haplotypes effects that may not be captured by a clump and threshold approach are *APOE* (associated with Alzheimer’s disease) (7) and *APOL1* (associated with chronic kidney disease) (8) haplotypes. Many other haplotypes with lower effect sizes may be yet unknown and harder to detect. In addition, clumping may not select causal variants or optimally tag SNPs for the population at hand. Moreover, effect sizes based on summary statistics from a GWAS conducted in one population may not be optimal for a different population. Specifically, PRS performance is known to be significantly affected by the population in which the GWAS was conducted, and PRS may not generalize well to different populations (9–11).

Some of the challenges of PRS modeling can be addressed using advanced machine learning (ML) methodologies. Many ML algorithms such as random forests, gradient boosted trees, and neural networks are explicitly non-linear, and allow interaction between features. Gradient boosted trees, for example, allow for the effect size of a given SNP to vary depending on the presence of an allele of a different SNP (12). Accordingly, ML methods have been used successfully to improve the prediction of complex phenotypes using genetic data (13). For example, a study employing random forests to predict type 2 diabetes found that it out-performed linear models, such as support vector machines (14). Gradient boosted trees have been used to predict breast cancer risk (15). Another study, however, have found that ML models that explicitly attempt to model non-linear effects do not perform as well as standard PRS (16). Finally, while ML models do not explicitly allow for generalization to non-sampled populations, we hypothesize that a large and ancestry diverse cohort would improve genetic prediction across populations.

Here, we explore the use of genetic data in prediction of five complex phenotypes: three established cardiovascular disease risk factors (total cholesterol levels (17), triglycerides (18), and systolic blood pressure (19)), sleep duration, a phenotype of lower heritability that is also associated with cardiovascular disease (20,21), and height, a highly heritable and well-studied phenotype. For each of these five complex phenotypes, we develop ensemble machine learning models for genetic trait prediction accounting for interactions, trained on a multi-ethnic dataset from the National Heart Lung and Blood Institute’s Trans-Omics in Precision Medicine (TOPMed) consortium (22). We examine the accuracy of the results to clump and threshold (C+T) PRS and explicitly compare ML models that allow for interactions and non-linear effects to those that do not. Finally, we assess the accuracy of the predictions for the ML models and the PRS models among White, Black, and Hispanic/Latino race/ethnic groups.

## Methods

### Study population

The study sample included 34,072 unrelated (3^rd^ degree or less) TOPMed participants from eight U.S. based cohort studies: Jackson Heart Study (JHS; n = 2,504), Framingham Heart Study (FHS; n = 3,520), Hispanic Community Health Study/Study of Latinos (HCHS/SOL; n=6,408), Atherosclerosis Risk in Communities study (ARIC; n=6,197), Cardiovascular Health Study (CHS; n=2,835), Multi-Ethnic Study of Atherosclerosis (MESA; n=3,949), Cleveland Family Study (CFS; n=1,182), and Coronary Artery Risk Development in Young Adults Study (CARDIA; n=2,468). Study descriptions are provided in Supplementary Materials. Phenotypes were harmonized by the TOPMed Data Coordinating Center (DCC) (23), and included age, sex, race/ethnicity, study (used as covariates), phenotypes of interest, and medications, which were used to adjust measures of relevant phenotypes (Supplementary Table S1). The dataset included 7,601 non-Hispanic Black participants, 14,142 non-Hispanic White participants, and 7,320 participants of Hispanic/Latino descent. All participants provided informed consent, and the study was approved by IRBs in each of the participating institutions. The dataset was divided such that 20% of the data was held out as a validation set. A secondary analysis used a larger training dataset that included related individuals but in which all individuals in the training dataset were still unrelated to those in the test dataset.

### Genotype data

We used whole genome sequencing data from TOPMed (24) Freeze 8, without restriction on sequencing depth, which contains 705,486,649 variants. The dataset includes samples sequenced through the National Human Genome Research Institute’s Centers for Common Disease Genomics (CCDG) program, where the sequence data for all TOPMed and CCDG samples were harmonized together via joint allele calling. The methods for TOPMed WGS data acquisition and quality control (QC) are provided in https://www.nhlbiwgs.org/topmed-whole-genome-sequencing-methods-freeze-8. Principal Components (PCs) and kinship coefficients were computed for the genetic data by the TOPMed DCC using the PC-Relate algorithm (25) implemented in the GENESIS R package (26). In this work, we used 5 PCs computed via the GENESIS R package PC-Air algorithm (27) to adjust for global ancestry. Based on the kinship coefficients, we identified related individuals and generated a dataset in which all individuals were degree-3 unrelated, i.e., all kinship coefficients were lower than 0.0625. We extracted allele counts of variants that passed QC from GDS files using the SeqArray (28) package version 1.28.1 and then further processed using R and Python scripts. For all variants, we set the effect allele to be the minor allele.

### Heritability estimation

Let **K** denote an *n* × *n* kinship matrix, having twice the kinship coefficient between the *i-th* and *j-th* participants in its *i,j* entry. For an outcome *y*, we assume the linear model:

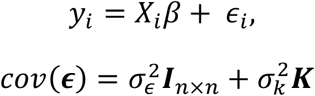

where ***ϵ*** = (*ϵ*_1_, …, *ϵ*_*n*_)^*T*^ is the normally-distributed vector of errors across the sample. We estimated the narrow-sense heritability 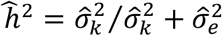 using the Restricted Maximum Likelihood approach as implemented in the GCTA (29) software (version 1.93.2).

## Phenotypes

We trained genetic prediction models to predict height, systolic blood pressure, total cholesterol, triglycerides concentration, and self-reported sleep duration, and used sex, study, race/ethnicity, and age as covariates. For reproducibility, Supplementary Table S1 provides the coded names of each of the phenotypes and covariates used in the analysis.

For each of the covariate-adjusted phenotypes of interest, we excluded outlying individuals defined by phenotypic values above the 99^th^ quantile and values below the 1^st^ quantile for the phenotype, computed over the multi-ethnic dataset. Then, each phenotype was regressed on age, sex, study, and race/ethnicity. The residuals were extracted and rank-normalized. Subsequent analyses used these rank-normalized residuals as the outcomes (30), and we refer to them henceforth as “adjusted phenotypes”.

### Summary statistics from published GWAS

We used summary statistics from published GWAS to select SNPs and their weights to construct PRS, as well as to select SNPs to include in the ML models. The GWAS used for each of the five phenotypes are described in Table 1. When possible, we used multi-ethnic GWAS. We lifted over the coordinates to our genome build GRCh38/hg38 using the LiftOver tool from the UCSC genome browser (31).

**Table 1.** Description of published GWAS used to identify summary statistics. GWAS source, study population as reported by the manuscript reporting the GWAS, and number of participants used to generate summary statistics for a given phenotype.

| *Phenotype* | *GWAS* | *Study Population* | *Participants* |
| --- | --- | --- | --- |
| **Height** | Meta-analysis of genome-wide association studies for height and body mass index in ~700000 individuals of European ancestry (32) | UK Biobank and the GIANT consortium | 693,529 (European ancestry) |
| **Total Cholesterol** | Genetics of Blood Lipids Among ~300,000 Multi-Ethnic Participants of the Million Veteran Program (33) | Million Veteran Program | 297,626 (72.4% non-Hispanic Whites, 19.3% non-Hispanic Blacks, 8.3% Hispanics) |
| **Triglycerides** | Genetics of Blood Lipids Among ~300,000 Multi-Ethnic Participants of the Million Veteran Program (33) | Million Veteran Program | 291,933 (72.4% non-Hispanic Whites, 19.3% non-Hispanic Blacks, 8.2% Hispanics) |
| **Systolic Blood Pressure** | Trans-ethnic association study of blood pressure determinants in over 750,000 individuals (34) | Million Veteran Program | 318,891 (69.1% non-Hispanic Whites, 18.8% non-Hispanic Blacks, 6.7% Hispanics, 0.77% non-Hispanic Asians and 0.85% non-Hispanic Native Americans) |
| **Sleep Duration** | Genome-wide association study identifies genetic loci for self-reported habitual sleep duration supported by accelerometer-derived estimates (35) | UK Biobank | 446,118 (European ancestry) |

### Polygenic Risk Score

We calculated the standard PRS using the classic clump-and-threshold methodology (C+T PRS) (3). We used PRSice 2 software version 2.3.1 (36) to calculate the genetic score with a clumping region of 250 kb on each side of the index SNP and a clumping R^2^ of 0.1. We considered p-value thresholds of 0.5 through 1e^-10^. For each adjusted phenotype we fit a linear model including covariates, the PRS, and genetic PCs to account for population structure (37) and selected the PRS in which the PRS model minimized the mean squared error in the training dataset. We assessed the percentage of variance explained (PVE) by the PRS models using the methodology described below.

### LASSO and Gradient Boosted Trees (XGBoost) ensemble

Figure 1 describes the construction of an ensemble ML model for polygenic risk prediction. We considered for inclusion in the models all SNPs having p-value<1×10^−4^ in the corresponding GWAS and used them to develop an ensemble prediction model. In brief, the ensemble model included two steps: (1) a LASSO penalized regression for filtering candidate SNPs; and (2) an XGBoost prediction model allowing for non-linear interactions. In detail, gradient boosted trees are a widely used machine learning technique that creates an ensemble of “weak” decision trees (i.e., limited in depth or interactions) by iteratively optimizing an objective function at each boosting step in which new trees are optimized based on the residuals of the previous boosting step. XGBoost is an optimized implementation of gradient boosted trees that is highly efficient in distributed computing environments (12). However, the set of candidate SNPs is very large for most of the GWAS listed in Table 1, and boosting is prone to overfitting with high dimensionality (38). LASSO (39) is a commonly used model for feature selection that can mitigate overfitting by encouraging parsimony through L1 regularization. We trained an ensemble model, jointly training LASSO and XGBoost models in order to prevent overfitting due to the high dimensionality of genetic data (through LASSO) while simultaneously exploiting the non-linear relationships and interaction effects (through XGBoost).

**Figure 1:**
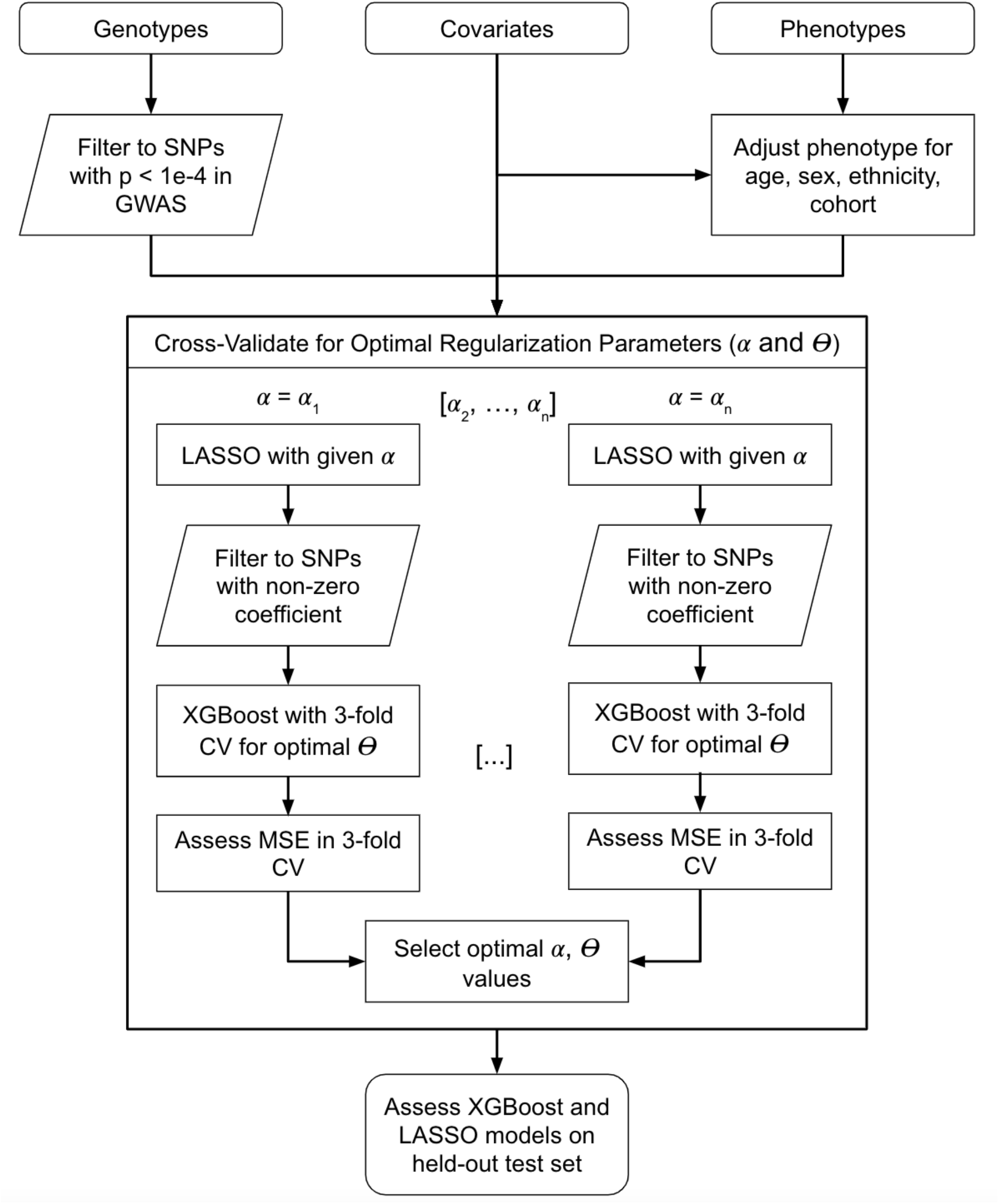
Flow chart of ensemble model structure. The model relies on jointly training the LASSO and XGBoost model to identify the optimal value for the L1 regularization parameter and the number of boosting steps. “CV” indicates cross-validation, *α* refers to the regularization parameter, and ⊖ is the number of boosted trees for XGBoost. The optimal values for these hyperparameters were selected using 3-fold CV for the mean squared error of the XGBoost model.

The ensemble model was trained as follows. For each given regularization hyperparameter *α* ∈ {0 … 1} we fit LASSO on the training dataset using a 10-fold cross validation scheme (and the MSE loss) and filtered to SNPs with non-zero coefficients from the LASSO model. The LASSO model included only linear SNP effects and unpenalized covariates. Using the selected SNPs, we fit the XGBoost model via 3-fold cross-validation applied on the training dataset, allowing up to 10,000 boosted trees with early stopping after 10 rounds of boosting without improvement in the 3-fold cross-validation loss (see Table 3 for details). Based on this 3-fold cross validation, we selected the number of trees *θ*_*α*_ that minimized the mean squared prediction error (MSE), resulting in a set of parameters (X, *θ*_*α*_). We selected the optimal (X, *θ*_*α*_) pair that minimized the MSE of the 3-fold cross validation step across all values of X. For XGBoost, we always used a learning rate of 0.01, maximum depth of 5, column sample by tree of 90%, minimum child weight of 10, and subsample of 50%. Finally, we performed LASSO regression using the same variants that were selected in this process, to explicitly compare the results of a non-linear model allowing for interactions to a linear model without interactions. Separately, we performed 3-fold cross-validation to select the optimal regularization hyperparameter with respect to the LASSO model in a separate process (Table S4).

We performed this process individually for each of our five adjusted phenotypes using a distributed cluster computing environment. All models included the ancestral PCs, and some models used the C+T PRS as a variable. We assessed the PVE of the genetic machine learning models using the methodology described below and compared the results with the standard PRS model. Analysis was conducted using Python 3 and the *scikit-learn* (40) *and xgboost* packages (12).

### SNP selection via joint LASSO/XGBoost hyperparameter tuning scheme

### Race/Ethnicity Analysis

We first trained the models using the combined, multi-ethnic dataset (“multi-ethnic model”). We then trained the models on the subset of the sample containing only White, Black, and Hispanic/Latino participants. This resulted in four models that were each trained on different race/ethnicity groups: Multi-Ethnic, White, Black, and Hispanic/Latino. For each of these four models, we assessed the PVE among the participants of each race/ethnicity in the held-out test set.

### Model evaluation in the held-out test set

We quantify model performance as the variance explained. Let 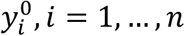 denote the adjusted phenotype. Var(*y*^0^) estimates the total baseline model variance. For a given model *m*, let 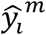 denote the predicted (adjusted) phenotype value for the *i*th person. We estimate the percent variance explained by model *m* as:

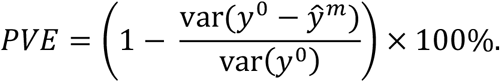

## Results

We used a multi-ethnic dataset from TOPMed containing 29,063 genotyped individuals from eight distinct cohorts (JHS, FHS, HCHS/SOL, ARIC, CHS, MESA, CFS and CARDIA) to train non-linear polygenic risk prediction models in diverse populations for five complex phenotypes: triglycerides, total cholesterol, systolic blood pressure, sleep duration, and height. Table 2 characterizes these phenotypes and covariates across the pooled and race/ethnicity-stratified training dataset based on unrelated individuals. We evaluate the models on an independent test dataset of 5,009 individuals (Supplementary Table S3). Results based on a training dataset that included related individuals (and still being unrelated to the test dataset were similar and are provided in the Supplementary Materials.

**Table 2.** Summary statistics of phenotypes used in the training dataset. Mean, Median and percent of missing data for the phenotypes (Triglycerides, Total Cholesterol, Systolic Blood Pressure, Sleep Duration and Height) and covariates (sex and age) used in this study. All the phenotypes are presented for the whole database as well as stratified by race/ethnicity (Black, White, and Hispanic/Latino). Summary statistics for the test dataset are provided in Supplementary Table S3.

|  | Black<br>(N=7601) | Hispanic/Latino<br>(N=7320) | White<br>(N=14142) | Overall<br>(N=29063) |
| --- | --- | --- | --- | --- |
| **Sex** |  |  |  |  |
| Male | 3066 (40.3%) | 3088 (42.2%) | 6432 (45.5%) | 12586 (43.3%) |
| Female | 4535 (59.7%) | 4232 (57.8%) | 7710 (54.5%) | 16477 (56.7%) |
| **Age** |  |  |  |  |
| Mean (SD) | 50.6 (16.9) | 48.2 (14.3) | 50.2 (16.4) | 49.8 (16.1) |
| Median [Min, Max] | 52.0 [2.00, 93.0] | 49.0 [5.00, 86.0] | 51.0 [3.00, 98.0] | 51.0 [2.00, 98.0] |
| **Triglycerides** |  |  |  |  |
| Mean (SD) | 106 (69.1) | 135 (96.0) | 125 (82.2) | 124 (84.2) |
| Median [Min, Max] | 90.0 [16.0, 1930] | 113 [20.0, 1670] | 106 [17.0, 1600] | 103 [16.0, 1930] |
| Missing | 2598 (34.2%) | 1316 (18.0%) | 3073 (21.7%) | 6987 (24.0%) |
| **Total Cholesterol** |  |  |  |  |
| Mean (SD) | 198 (41.8) | 200 (43.2) | 205 (39.2) | 202 (41.0) |
| Median [Min, Max] | 196 [74.0, 450] | 197 [62.0, 526] | 202 [77.8, 594] | 199 [62.0, 594] |
| Missing | 2598 (34.2%) | 1316 (18.0%) | 3073 (21.7%) | 6987 (24.0%) |
| **Systolic Blood Pressure** |  |  |  |  |
| Mean (SD) | 127 (20.9) | 121 (17.2) | 118 (17.1) | 121 (18.5) |
| Median [Min, Max] | 123 [73.0, 246] | 119 [77.0, 218] | 116 [67.0, 227] | 118 [67.0, 246] |
| Missing | 1944 (25.6%) | 1589 (21.7%) | 2972 (21.0%) | 6505 (22.4%) |
| **Sleep Duration** |  |  |  |  |
| Mean (SD) | 6.50 (1.51) | 7.73 (1.52) | 7.09 (1.16) | 7.15 (1.44) |
| Median [Min, Max] | 6.00 [1.00, 16.5] | 7.79 [2.00, 13.4] | 7.00 [1.00, 16.0] | 7.00 [1.00, 16.5] |
| Missing | 2352 (30.9%) | 411 (5.6%) | 4468 (31.6%) | 7231 (24.9%) |
| **Height** |  |  |  |  |
| Mean (SD) | 168 (10.4) | 163 (9.24) | 168 (10.3) | 167 (10.3) |
| Median [Min, Max] | 168 [85.7, 207] | 162 [116, 194] | 168 [94.0, 203] | 166 [85.7, 207] |

### XGBoost outperforms linear models for the prediction of complex phenotypes

We constructed three models of increasing complexity for each of the five phenotypes (Figure 1). Each phenotype was adjusted for the covariates and the residuals were rank-normalized and standardized. Each model was then fine-tuned to predict the residuals from genetic SNP data. The three models employed in this study were C+T PRS, LASSO, and XGBoost, with the number of SNPs selected for each algorithm for each phenotype listed in Table 3. The hyperparameters selected for each model through cross-validation are listed in Table S8.

**Table 3.** Number of SNPs selected through cross-validation. Displayed are number of SNPs selected for each of the phenotypes in the four models in this study: C+T PRS, XGBoost alone, LASSO (which has the same number of variants as in the “XGBoost alone” model, because the LASSO selected the variants used by XGBoost), and XGBoost with C+T PRS (for this model we do not additionally report results from a model based on LASSO alone using the same variants).

| Phenotype | C+T PRS | XGBoost alone | Lasso | XGBoost with PRS |
| --- | --- | --- | --- | --- |
| Triglycerides | 7500 | 432 | 432 | 304 |
| Total Cholesterol | 6960 | 455 | 455 | 455 |
| Systolic Blood Pressure | 8066 | 72 | 72 | 346 |
| Sleep Duration | 4876 | 91 | 91 | 666 |
| Height | 22461 | 2193 | 2193 | 916 |

Figure 2 depicts the PVE across different prediction models and phenotypes. The C+T PRS outperformed the LASSO model, except for total cholesterol, even though the LASSO model used only 6.5% of the SNPs used by the C+T PRS. The XGBoost algorithm trained directly on SNPs (“XGBoost alone”), outperforms linear models for both total cholesterol and triglycerides. For height, it significantly underperformed the C+T PRS model.

**Figure 2:**
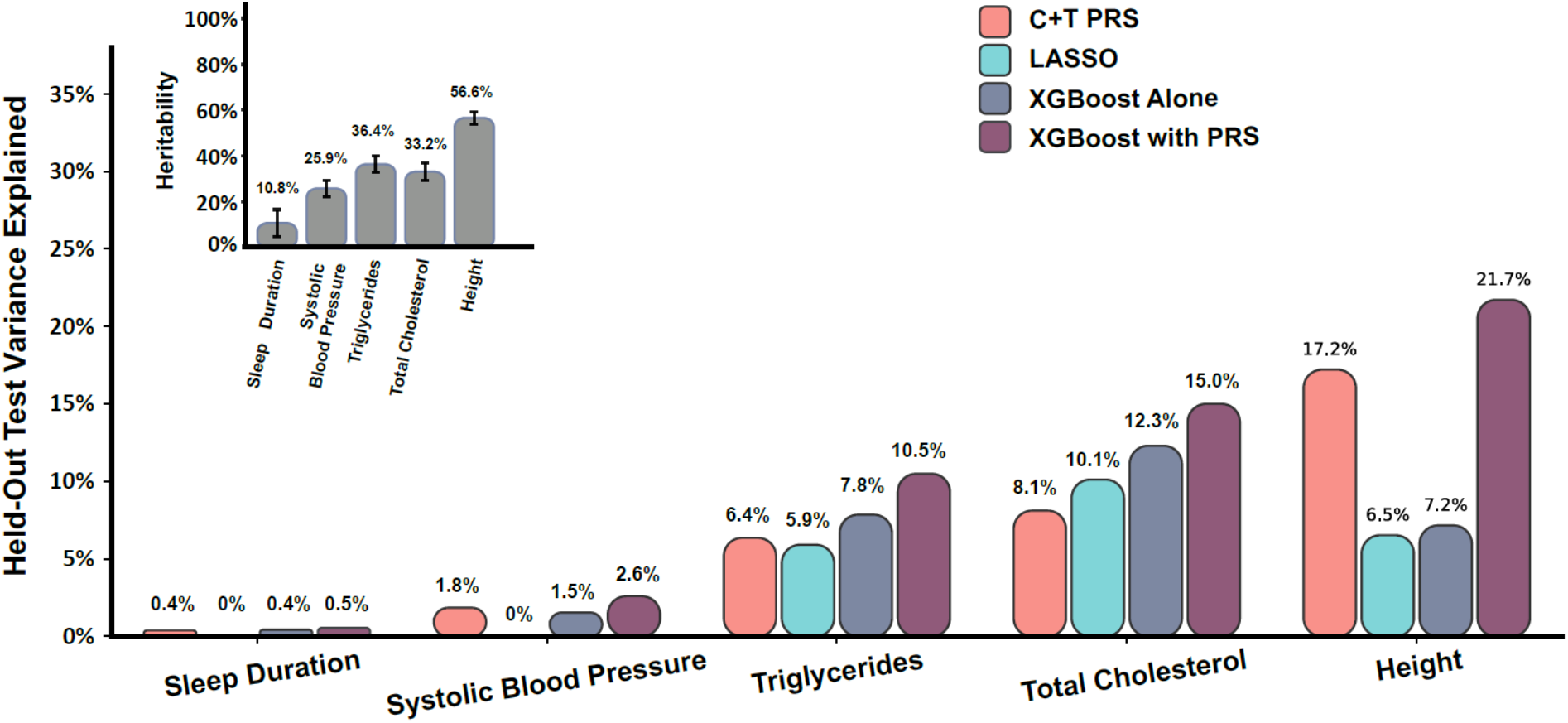
Non-linear model consistently outperforms linear ones for prediction of multiple complex phenotypes in multi-ethnic dataset. Linear (C+T PRS-pink), linear-regularized (LASSO – teal), and non-linear (XGBoost – grey, purple) models were employed to predict the harmonized triglycerides, total cholesterol, systolic blood pressure, sleep duration and height phenotypes from SNP data from TOPMed following adjustment for covariates. Two versions of the XGBoost algorithm are shown with the first model employing only the SNPs as features (grey; “XGBoost alone”) and a second model which had the C+T PRS as one of the features as well (“XGBoost with PRS”). The LASSO algorithm (teal) was trained on the same set of SNPs as the XGBoost. The inset depicts estimated heritabilities for same phenotypes in the same database using the REML approach.

### Modeling of non-linearities and interactions among SNPs improves the prediction of complex human phenotypes

The non-linear XGBoost algorithm outperforms the linear LASSO when trained on the same SNP set (Figure 2 grey vs teal) for all phenotypes. The improved performance may stem either from modelling non-linear genetic effects or interactions between SNPs, or both, since both of these are addressed by the algorithm (12). However, both XGBoost and the LASSO PRS sometimes underperformed the C+T PRS, likely because the C+T PRS was able to combine information from multiple SNPs. Notably, for height, the XGBoost model had only 7.2 PVE while the C+T PRS had 17.2 PVE. This could also be due to overfitting to the training dataset. To combine the advantages of a genome-wide PRS and of the XGBoost accounting for non-linearities and interactions, we constructed an additional, “XGBoost with PRS”, model that included both the individual SNPs as described in Figure 1, and also the C+T PRS score. Indeed, we see that this model substantially outperforms the linear C+T and LASSO models, as well as XGBoost alone, for all phenotypes, providing a strong indication for non-linear effects and/or genotype by genotype interactions. Substantial improvement is observed for all phenotypes. Specifically, compared to the C+T PRS baseline, the XGBoost with C+T PRS improved the PVE by 64% for triglycerides, 85% for total cholesterol, 44% for systolic blood pressure, 25% for sleep duration, and 26% for height.

Notably, even our best model falls short compared to the estimated heritability obtained from a linear mixed model that considers all SNPs via the kinship matrix (Figure 2 inset). For example, we achieved ∼2.5-fold better results for height (21.7% vs 56.6%) and ∼10-fold better results for systolic blood pressure (2.6% vs 25.9%) with linear mixed models. These results indicate that much of the effect is distributed among a large number of weakly-correlated SNPs.

### Race/Ethnicity associates with model performance for multiple phenotypes

Our dataset included participants with self-reported race/ethnicity (7601 Black, 14142 White, and 7320 Hispanic/Latino), with phenotype characteristics provided in Table 2. We compared the performance of the simple C+T PRS model with the best-performing XGBoost model that includes the C+T PRS as a feature, trained on the combined dataset, for the prediction of the different phenotypes on the ethnicity-specific datasets (Figure 3).

**Figure 3:**
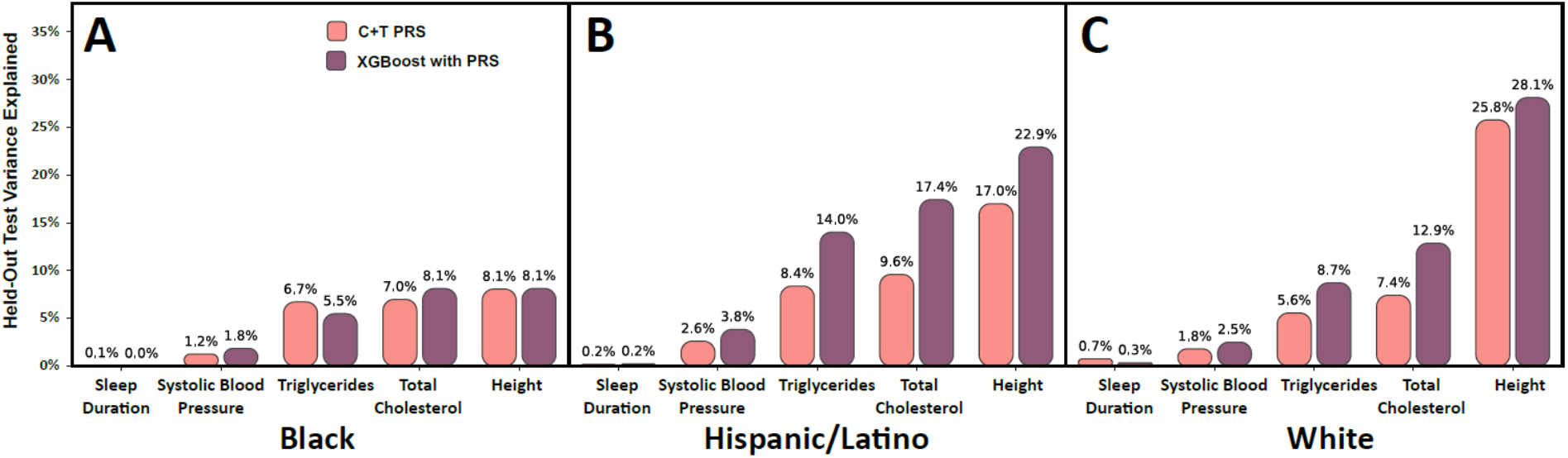
Model performance differ by group, with XGBoost consistently outperforming PRS. Performance of the C+T PRS (pink) and XGBoost+PRS (purple) models trained on the combined dataset when applied to the prediction of the 5 phenotypes in separate race/ethnicities. **(A**,**B**,**C)** White, Black and Hispanic/Latino groups accordingly.

The XGBoost with PRS model usually improves PVEs over the C+T PRS in the White and Hispanic/Latino groups, but less so in the Black group. Surprisingly, for a few phenotypes (systolic blood pressure, triglycerides, and total cholesterol), the PVEs were better in Hispanics/Latinos compared to Whites, even though the GWAS using from more data from Whites than Hispanic/Latino participants. However, it is important to note that many Hispanic/Latino individuals have substantial European genetic ancestry, and our study does not differentiate between Hispanic/Latinos with different levels of European genetic ancestry. Unfortunately, most models performed poorly in Blacks.

### Ethnic diversity is crucial for model training

Figure 4 compares XBoost models trained within race/ethnic group to the multi-ethnic model.

**Figure 4:**
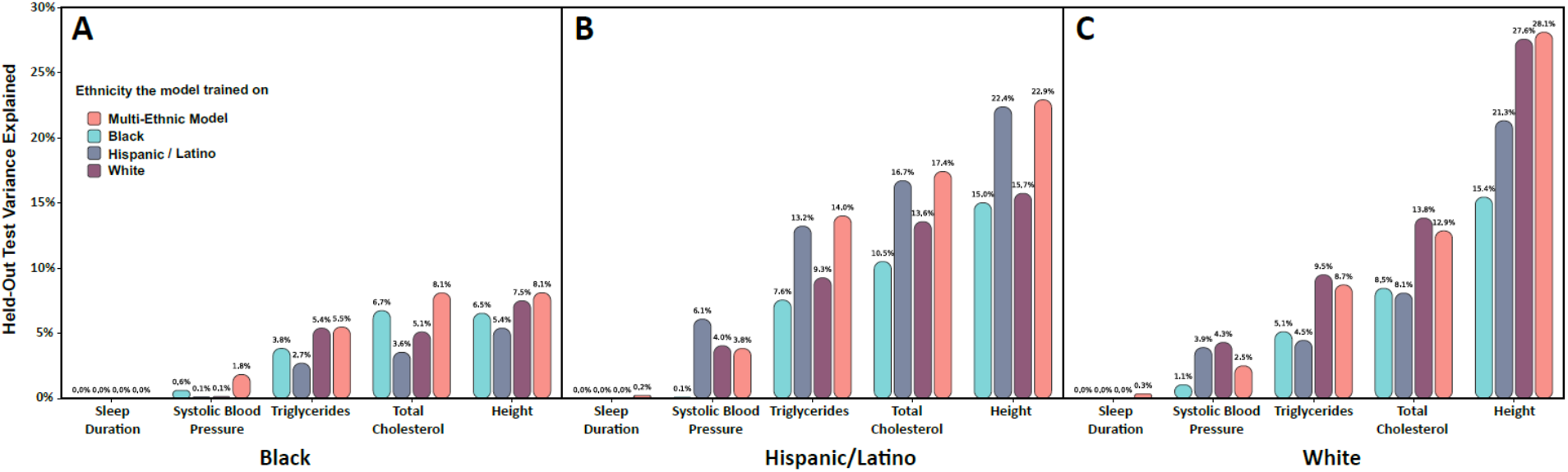
Multi-ethnic XGBoost model performs on par with the race/ethnic-specific models. XGBoost with PRS models (see Materials and Methods) were trained either on the combined dataset containing all participants, (pink) or on each race/ethnic group separately (teal, grey and purple). The models were then evaluated on each of the groups (left panel - Black, middle panel - Hispanic/Latino, and right panel - White).

For the Black group, the multi-ethnic model performed best on the held-out test datasets, consistently outperforming the race/ethnic specific models – even those trained and tested on the same race/ethnic group. However, for the Hispanic/Latino group, the multi-ethnic model was sometimes inferior to the race/ethnic matched model (for systolic blood pressure). For Whites, the race/ethnic matched model improves upon the multi-ethnic model for almost all phenotypes (except height and sleep duration). This may be due to the larger sample size available in the multi-ethnic training dataset, compared to the Black and Hispanic/Latino datasets.

## Discussion

The aim of this study was to investigate the application of machine learning algorithms to polygenic trait prediction, specifically algorithms that allow for non-linearities and interaction effects between SNPs, and to compare their performance to standard C+T PRS and LASSO methodologies that do not account for such effects. We chose five complex phenotypes with varying levels of heritability across a large, multi-ethnic dataset including White, Black, and Hispanic/Latino participants.

Across all phenotypes in the validation dataset, we found the highest PVE by combining the XGBoost model with the C+T PRS. The increase in PVE varied across the phenotypes, with up to 85% for total cholesterol. The impressive increase in the performance of the XGBoost model relative to C+T PRS and LASSO points toward interactions between genetic alleles and/or non-linear contributions of SNPs to phenotypes. In almost all cases, the XGBoost algorithm alone (without including the C+T PRS score) out-performed the linear LASSO model that used exactly the same SNPs. In most cases, however, the C+T PRS performed better, likely because it could account for more weakly-associated SNPs. Combining the ML model with the C+T PRS (as a feature) achieved high prediction performance by both accounting for the large numbers of weakly associated SNPs (linearly through C+T PRS), in addition to some of the non-linearities and interactions (through XGBoost).

We chose to employ the XGBoost implementation of gradient boosted trees due to its strong performance in prediction tasks, explicit handling of interactions, and ability to capture non-linear effects. The large number of potential SNPs precluded their direct inclusion into the XGBoost model, as it is extremely computationally expensive and prone to overfitting with high dimensionality. Thus, we developed an ensemble model that used the LASSO algorithm as a feature selection tool to optimize the XGBoost performance while performing a cross-validation for the hyperparameters of both LASSO and XGBoost. The inclusion of both C+T PRS scores as well as XGBoost allows for direct comparison between the models, with the C+T PRS representing the linear additive genetic contributions to the trait, allowing XGBoost to optimize the non-linear and interaction effects.

Several studies that compared linear effects PRS models to ML models allowing for more complex genetic effects reported that standard PRS outperforms ML models. For example, one study suggested that a PRS for coronary artery disease outperformed a variety of ML models (16). Another study found that a linear Elastic Net model usually outperformed ML models that allow for non-linear and interaction effects for prediction of gene transcripts (41). And yet another study found that linear PRS models outperformed support vector machines for psychiatric phenotypes (42). Our study differs from these prior studies by use of very large and diverse training and testing datasets (prior studies were often limited to a few thousand individuals). Our datasets also had high quality deep sequencing and joint allele calling, as well as harmonized phenotypes across the combined dataset, which likely improved our ability to validate ML models across training and testing datasets. Also, our novel ensemble approach to guarding against overfitting, in addition to including the standard linear model PRS within the XGBoost model, us utilized the strengths of both the linear and the non-linear approaches in complementary ways. Specifically, this approach leveraged the ability of the PRS to capture the linear additive effects from a large number of SNPs, and the XGBoost to capture non-linear effects and SNP-SNP interactions.

We compared multi-ethnic and race/ethnicity-specific models and found that multi-ethnic-trained models on large datasets had different performance across race/ethnic groups. In our analyses, the multi-ethnic models had better performance in Whites and the Hispanic/Latino group than in Blacks. Models trained using the same race/ethnic group and the multi-ethnic trained model had similar prediction performance, despite a substantial decrease in training sample size in the multi-ethnic model. Overall, we found that the PVE for the studied phenotypes was consistently lower for Black participants than for White or Hispanic/Latino participants. The difference in PVE varied by phenotype, from 1.3 to 3.5 times lower for Black participants compared to White. There are several possible explanations for these findings. First it may be that the combined models predominately use European-ancestry specific genetic effects. Both the White and the Hispanic/Latino groups have substantial European ancestry, while Black groups have lower European ancestry. Specifically, across Hispanic/Latino background groups reported in the Hispanic Community Health Study/Study of Latinos, on average 40-80% have European ancestry (43) while in the Jackson Heart Study, Blacks are estimated to have. 16% European ancestry on average (44). For three of the phenotypes we used multi-ethnic GWAS analyses to select candidate SNPs for analysis. However, most GWAS participants are still White (45). Therefore, the choice of SNPs is more optimal for groups with substantial amount of European ancestry, so that SNPs with small effects or low minor allele frequencies (MAF) in European ancestry populations and larger effects or higher MAF in African ancestry populations were not discovered in the GWAS and were therefore not selected to be used in the trained prediction models. This limitation has been shown to reduce PRS performance in African and African Americans populations in multiple studies (46–48).

There are some limitations to this study. First, while the TOPMed cohort is diverse, White participants are over-represented. Second, as noted above, although the GWAS analyses that we relied upon were multi-ethnic (other than sleep duration and height GWAS which were based European ancestry samples), it seems likely that important variants for these phenotypes among a Black population do not achieve the required p-value level (<10^−4^) to be included, given limited sample sizes for Black participants in these prior GWAS analyses. Third, much of our ensemble algorithm relies on feature selection. This may be overly restrictive and does not allow for variants with very small effect sizes to be included (as noted in the results for Height). Fourth, we used self-reported race/ethnicity. An alternative grouping would use genetically-determined ancestry groups. We chose self-reported grouping to better approximate clinical settings and to potentially account for gene-environment interactions, in which people who share self-reported race/ethnicity may have more similar environmental exposures, compared to individuals outside the group. Fifth, we use ML as a tool to model the interactions and non-linearities. However, this approach does not explicitly identify individual interactions or non-linearities nor quantifies the contributions of each.

Overall, this study uncovers strong evidence for contributions of non-linear genetic effects and interaction between alleles to complex phenotypes. Additionally, our findings re-iterate one of the largest hurdles for better performing, robust genetic prediction models across diverse individuals – namely the lack of well-powered GWAS for different race/ethnic groups and subpopulations (49). This work opens up promising avenues for future research, such as: creating a generalizable tool that would allow ML PRS to be deployed on other studies; estimating the individual contributions of interactions and non-linearities; and developing approaches to prioritize SNPs for inclusion in the ML model that would increase predictive ability in Black and other non-White populations.

## Data Availability

TOPMed freeze 8 WGS data are available by application to dbGaP according to the study specific accessions: FHS: phs000974.v4.p3, JHS: phs000964.v1.p1, MESA: phs001211.v3.p2, CARDIA: phs001612.v1.p1, CFS: phs000954.v3.p2, CHS: phs001368.v2.p1, HCHS/SOL: phs001395.v1.p1, ARIC phs001416.v2.p1. Study phenotypes are available from dbGaP from parent studies accession: FHS: phs000007.v32.p13, JHS: phs000286.v6.p2, MESA: phs000209.v13.p3, CARDIA: phs000285.v3.p2, CFS: phs000284.v2.p1, CHS: phs000287.v7.p1, HCHS/SOL: phs000810.v1.p1, ARIC: phs000090.v7.p1.

https://dbgap.ncbi.nlm.nih.gov/

## Description of Supplemental Data

The Supplemental Data provides description of all TOPMed studies participating in the present analysis, including study-specific acknowledgements and ethics statements about informed consent and Institutional Review Boards approving each study. It also provides additional information on phenotypes used in the analyses, tables and figures.

## Acknowledgements

Molecular data for the Trans-Omics in Precision Medicine (TOPMed) program was supported by the National Heart, Lung and Blood Institute (NHLBI). Genome sequencing for “NHLBI TOPMed: Whole Genome Sequencing and Related Phenotypes in the Framingham Heart Study” (phs000974.v4.p3) was performed at the Broad Institute Genomics Platform (3R01HL092577-06S1, 3U54HG003067-12S2). Genome sequencing for “NHLBI TOPMed: The Jackson Heart Study” (phs000964.v1.p1) was performed at the Northwest Genomics Center (HHSN268201100037C). Genome sequencing for the “NHLBI TOPMed: The Atherosclerosis Risk in Communities Study” (phs001211.v3.p2) was performed at the Broad Institute Genomics Platform (3R01HL092577-06S1) and the Baylor College of Medicine Human Genome Sequencing Center (HHSN268201500015C, 3U54HG003273-12S2). Genome sequencing for “NHLBI TOPMed: Coronary Artery Risk Development in Young Adults Study” (phs001612.v1.p1) was performed at the Baylor College of Medicine Human Genome Sequencing Center (HHSN268201600033I). Genome sequencing for “NHLBI TOPMed: Cleveland Family Study” (phs000954.v3.p2) was performed at the Northwest Genomics Center (3R01HL098433-05S1, HHSN268201600032I). Genomics sequencing for “NHLBI TOPMed: Cardiovascular Health Study” (phs001368.v2.p1) was performed at the Baylor College of Medicine Human Genome Sequencing Center (3U54HG003273-12S2, HHSN268201500015C, HHSN268201600033I). Genome sequencing for “NHLBI TOPMed: Hispanic Community Health Study/Study of Latinos” (phs001395.v1.p1) was performed at the Baylor College of Medicine Human Genome Sequencing Center (HHSN268201600033I). Genome sequencing for “NHLBI TOPMed: Multi-Ethnic Study of Atherosclerosis” (phs001416.v2.p1) was performed at Broad Institute Genomics Platform (HHSN268201500014C, 3U54HG003067-13S1). Core support including centralized genomic read mapping and genotype calling, along with variant quality metrics and filtering were provided by the TOPMed Informatics Research Center (3R01HL-117626-02S1; contract HHSN268201800002I). Core support including phenotype harmonization, data management, sample-identity QC, and general program coordination were provided by the TOPMed Data Coordinating Center (R01HL-120393; U01HL-120393; contract HHSN268201800001I). We gratefully acknowledge the studies and participants who provided biological samples and data for TOPMed. The Genome Sequencing Program (GSP) was funded by the National Human Genome Research Institute (NHGRI), the National Heart, Lung, and Blood Institute (NHLBI), and the National Eye Institute (NEI). The GSP Coordinating Center (U24 HG008956) contributed to cross-program scientific initiatives and provided logistical and general study coordination. The Centers for Common Disease Genomics (CCDG) program was supported by NHGRI and NHLBI, and whole genome sequencing was performed at the Baylor College of Medicine Human Genome Sequencing Center (UM1 HG008898 and R01HL059367). ME, TS, and SR are supported by NHLBI grants R35HL135818 to SR. GMP is supported by R01HL142711 from the NHLBI. The project described was supported by the National Center for Advancing Translational Sciences, National Institutes of Health, through grant KL2TR002490 to LMR. PdV was supported by American Heart Association grant number 18CDA34110116 and NHLBI grant number R01HL146860. The views expressed in this manuscript are those of the authors and do not necessarily represent the views of the National Heart, Lung, and Blood Institute; the National Institutes of Health; or the U.S. Department of Health and Human Services.

## Web Resources

Code used for the analyses in this manuscript is provided on a dedicated GitHub repository https://github.com/genevievelyons/MachineLearning_PolygenicRiskScore.

## Author contribution statement

ME, GL, SRB, and NK constructed PRS. ME and GL developed ML models, performed association analyses, and summarized results in tables and figures. ME, GL, and TS conceptualized and drafted the manuscript. TS supervised the work for this manuscript. AMS supervised the phenotype harmonization of the TOPMed DCC. JAB, XG, FL, LR, YG, LL, GMP, JIR, SSR, ACM, BMP, DL, and SL designed data collection and/or TOPMed sample selection in the cohort they represent, and designed best-practices for data analysis for the same cohorts.

## Consortia

A complete list of TOPMed consortium authors is appears in https://www.nhlbiwgs.org/topmed-banner-authorship.

## Description of studies participating in the analysis and their Ethics Statements

### HCHS/SOL

The Hispanic Community Health Study/Study of Latinos (dbGaP accession phs000810) is a community-based longitudinal cohort study of 16,415 self-identified Hispanic/Latino persons aged 18–74 years and selected from households in predefined census-block groups across four US field centers (in Chicago, Miami, the Bronx, and San Diego). The census-block groups were chosen to provide diversity among cohort participants with regard to socioeconomic status and national origin or background (1,2). The HCHS/SOL cohort includes participants who self-identified as having a Hispanic/Latino background; the largest groups are Central American (n = 1,730), Cuban (n = 2,348), Dominican (n = 1,460), Mexican (n = 6,471), Puerto Rican (n = 2,728), and South American (n = 1,068). The HCHS/SOL baseline clinical examination occurred between 2008 and 2011 and included comprehensive biological, behavioral, and sociodemographic assessments. Visit 2 took place between 2014 and 2017, which re-examined 11,623 participants from the baseline sample. Visit 3 has started in 2020 and will last 3 years. In addition to clinic visit, participants are contacted annually to assess clinical outcomes. The study was approved by the Institutional Review Boards at each participating institution and written informed consent was obtained from all participants.

#### Ethics statement

This study was approved by the institutional review boards (IRBs) at each field center, where all participants gave written informed consent, and by the Non-Biomedical IRB at the University of North Carolina at Chapel Hill, to the HCHS/SOL Data Coordinating Center. All IRBs approving the study are: Non-Biomedical IRB at the University of North Carolina at Chapel Hill. Chapel Hill, NC; Einstein IRB at the Albert Einstein College of Medicine of Yeshiva University. Bronx, NY; IRB at Office for the Protection of Research Subjects (OPRS), University of Illinois at Chicago. Chicago, IL; Human Subject Research Office, University of Miami. Miami, FL; Institutional Review Board of San Diego State University. San Diego, CA.

#### Acknowledgements

The Hispanic Community Health Study/Study of Latinos is a collaborative study supported by contracts from the National Heart, Lung, and Blood Institute (NHLBI) to the University of North Carolina (HHSN268201300001I / N01-HC-65233), University of Miami (HHSN268201300004I / N01-HC-65234), Albert Einstein College of Medicine (HHSN268201300002I / N01-HC-65235), University of Illinois at Chicago – HHSN268201300003I / N01-HC-65236 Northwestern Univ), and San Diego State University (HHSN268201300005I / N01-HC-65237). The following Institutes/Centers/Offices have contributed to the HCHS/SOL through a transfer of funds to the NHLBI: National Institute on Minority Health and Health Disparities, National Institute on Deafness and Other Communication Disorders, National Institute of Dental and Craniofacial Research, National Institute of Diabetes and Digestive and Kidney Diseases, National Institute of Neurological Disorders and Stroke, NIH Institution-Office of Dietary Supplements.

### FHS

The Framingham Heart Study (dbGaP accession phs000007) began in 1948 with the recruitment of an original cohort of 5,209 men and women (mean age 44 years; 55 percent women). In 1971 a second generation of study participants was enrolled; this cohort (mean age 37 years; 52% women) consisted of 5,124 children and spouses of children of the original cohort. A third-generation cohort of 4,095 children of offspring cohort participants (mean age 40 years; 53 percent women) was enrolled in 2002-2005 and are seen every 4 to 8 years. Details of study designs for the three cohorts are summarized elsewhere (3–5). At each clinic visit, a medical history was obtained, and participants underwent a physical examination. Only study participants consented for genetic and non-genetic data are included. FHS has been approved by the Boston University IRB.

#### Ethics statement

The Framingham Heart Study was approved by the Institutional Review Board of the Boston University Medical Center. All study participants provided written informed consent.

#### Acknowledgments

The Framingham Heart Study (FHS) acknowledges the support of contracts NO1-HC-25195, HHSN268201500001I and 75N92019D00031 from the National Heart, Lung and Blood Institute and grant supplement R01 HL092577-06S1 for this research. We also acknowledge the dedication of the FHS study participants without whom this research would not be possible. Dr. Vasan is supported in part by the Evans Medical Foundation and the Jay and Louis Coffman Endowment from the Department of Medicine, Boston University School of Medicine.

### ARIC

The Atherosclerosis Risk in Communities (ARIC) study (dbGaP accession phs000090) is a population-based prospective cohort study of cardiovascular disease sponsored by the NHLBI. ARIC included 15,792 individuals, predominantly European American and African American, aged 45-64 years at baseline (1987-89), chosen by probability sampling from four US communities. Cohort members completed three additional triennial follow-up examinations, a fifth exam in 2011-2013, a sixth exam in 2016-2017, and a seventh exam in 2018-2019. The ARIC study has been described in detail previously (6).

#### Ethics statement

The ARIC study has been approved by Institutional Review Boards (IRB) at all participating institutions: University of North Carolina at Chapel Hill IRB, Johns Hopkins University IRB, University of Minnesota IRB, and University of Mississippi Medical Center IRB. Study participants provided written informed consent at all study visits.

#### Acknowledgements

The Atherosclerosis Risk in Communities study has been funded in whole or in part with Federal funds from the National Heart, Lung, and Blood Institute, National Institutes of Health, Department of Health and Human Services (contract numbers HHSN268201700001I, HHSN268201700002I, HHSN268201700003I, HHSN268201700004I and HHSN268201700005I). The authors thank the staff and participants of the ARIC study for their important contributions.

### CHS

The Cardiovascular Health Study (dbGaP accession phs000287) is a population-based cohort study initiated by the NHLBI in 1987 to determine the risk factors for development and progression of cardiovascular disease (CVD) in older adults, with an emphasis on subclinical measures. The study recruited 5,888 adults aged 65 or older at entry in four U.S. communities and conducted extensive annual clinical exams between 1989-1999 along with semi-annual phone calls, events adjudication, and subsequent data analyses and publications. Additional data are collected by studies ancillary to CHS. In June 1990, four Field Centers (Sacramento, CA; Hagerstown, MD; Winston-Salem, NC; Pittsburgh, PA) completed the recruitment of 5201 participants. Between November 1992 and June 1993, an additional 687 African Americans were recruited using similar methods. Blood samples were drawn from all participants at their baseline examination and during follow-up clinic visits and DNA was subsequently extracted from available samples

#### Ethics statement

All CHS participants provided informed consent, and the study was approved by the Institutional Review Board [or ethics review committee] of University Washington.

#### Acknowledgements

The Cardiovascular Health Study was supported by contracts HHSN268201200036C, HHSN268200800007C, HHSN268201800001C, N01HC55222, N01HC85079, N01HC85080, N01HC85081, N01HC85082, N01HC85083, N01HC85086, 75N92021D00006, and grants U01HL080295 and U01HL130114 from the National Heart, Lung, and Blood Institute (NHLBI), with additional contribution from the National Institute of Neurological Disorders and Stroke (NINDS). Additional support was provided by R01AG023629 from the National Institute on Aging (NIA). A full list of principal CHS investigators and institutions can be found at CHS-NHLBI.org. The content is solely the responsibility of the authors and does not necessarily represent the official views of the National Institutes of Health.

### MESA

The Multi-Ethnic Study of Atherosclerosis (dbGaP accession phs000209) is a study of the characteristics of subclinical cardiovascular disease (disease detected non-invasively before it has produced clinical signs and symptoms) and the risk factors that predict progression to clinically overt cardiovascular disease or progression of the subclinical disease (7). MESA consisted of a diverse, population-based sample of an initial 6,814 asymptomatic men and women aged 45-84. 38 percent of the recruited participants were white, 28 percent African American, 22 percent Hispanic, and 12 percent Asian, predominantly of Chinese descent. Participants were recruited from six field centers across the United States: Wake Forest University, Columbia University, Johns Hopkins University, University of Minnesota, Northwestern University and University of California - Los Angeles. Participants are being followed for identification and characterization of cardiovascular disease events, including acute myocardial infarction and other forms of coronary heart disease (CHD), stroke, and congestive heart failure; for cardiovascular disease interventions; and for mortality. The first examination took place over two years, from July 2000 - July 2002. It was followed by five examination periods that were 17-20 months in length. Participants have been contacted every 9 to 12 months throughout the study to assess clinical morbidity and mortality.

### Ethics statement

All MESA participants provided written informed consent, and the study was approved by the Institutional Review Boards at The Lundquist Institute (formerly Los Angeles BioMedical Research Institute) at Harbor-UCLA Medical Center, University of Washington, Wake Forest School of Medicine, Northwestern University, University of Minnesota, Columbia University, and Johns Hopkins University.

#### Acknowledgments

The MESA projects are conducted and supported by the National Heart, Lung, and Blood Institute (NHLBI) in collaboration with MESA investigators. Support for MESA is provided by contracts 75N92020D00001, HHSN268201500003I, N01-HC-95159, 75N92020D00005, N01-HC-95160, 75N92020D00002, N01-HC-95161, 75N92020D00003, N01-HC-95162, 75N92020D00006, N01-HC-95163, 75N92020D00004, N01-HC-95164, 75N92020D00007, N01-HC-95165, N01-HC-95166, N01-HC-95167, N01-HC-95168, N01-HC-95169, UL1-TR-000040, UL1-TR-001079, and UL1-TR-001420. Also supported in part by the National Center for Advancing Translational Sciences, CTSI grant UL1TR001881, and the National Institute of Diabetes and Digestive and Kidney Disease Diabetes Research Center (DRC) grant DK063491 to the Southern California Diabetes Endocrinology Research Center.

### CARDIA

The Coronary Artery Risk Development in Young Adults study (dbGaP accession phs000285) is a prospective multicenter study with 5,115 adults Caucasian and African American participants of the age group 18-30 years at baseline, recruited from four centers at the baseline examination in 1985-1986 (8). The recruitment was done from the total community in Birmingham, AL, from selected census tracts in Chicago, IL and Minneapolis, MN; and from the Kaiser Permanente health plan membership in Oakland, CA. Nine examinations have been completed in the years 0, 2, 5, 7, 10, 15, 20, 25 and 30, with high retention rates (91%, 86%, 81%, 79%, 74%, 72%, 72%, and 71%, respectively) and written informed consent was obtained in each visit.

#### Ethics statement

All CARDIA participants provided informed consent, and the study was approved by the Institutional Review Boards of the University of Alabama at Birmingham and the University of Texas Health Science Center at Houston.

#### Acknowledgements

The Coronary Artery Risk Development in Young Adults Study (CARDIA) is conducted and supported by the National Heart, Lung, and Blood Institute (NHLBI) in collaboration with the University of Alabama at Birmingham (HHSN268201800005I & HHSN268201800007I), Northwestern University (HHSN268201800003I), University of Minnesota (HHSN268201800006I), and Kaiser Foundation Research Institute (HHSN268201800004I). CARDIA was also partially supported by the Intramural Research Program of the National Institute on Aging (NIA) and an intra-agency agreement between NIA and NHLBI (AG0005).

### JHS

The Jackson Heart Study (dbGaP accession phs000286) is a longitudinal investigation of genetic and environmental risk factors associated with the disproportionate burden of cardiovascular disease in African Americans (9,10). JHS is funded by the NHLBI and the National Institute on Minority Health and Health Disparities (NIMHD) and is an expansion of the ARIC study in its Jackson Field Center. At baseline, the JHS recruited 5306 African American residents of the Jackson Mississippi Metropolitan Statistical Area aged, approximately 6.6% of all African American adults aged 35-84 residing in the area. Participants were recruited via random sampling (17% of participants), volunteers (30%), prior participants in the Atherosclerosis Risk in Communities (ARIC) study (31%), and secondary family members (22%). Among these participants, approximately 3400 gave consent that allows genetic research. JHS has conducted three back-to-back clinical examinations (Exam 1, 2000-2004; Exam 2, 2005-2008; and Exam 3, 2009-2013), and a fourth clinical examination is underway. Participants are also contacted annually by telephone to update personal and health information including vital status, interim medical events, hospitalizations, functional status and sociocultural information.

#### Ethics statement

The JHS study was approved by Jackson State University, Tougaloo College, and the University of Mississippi Medical Center IRBs, and all participants provided written informed consent.

#### Acknowledgements and Disclaimer

The Jackson Heart Study (JHS) is supported and conducted in collaboration with Jackson State University (HHSN268201800013I), Tougaloo College (HHSN268201800014I), the Mississippi State Department of Health (HHSN268201800015I) and the University of Mississippi Medical Center (HHSN268201800010I, HHSN268201800011I and HHSN268201800012I) contracts from the National Heart, Lung, and Blood Institute (NHLBI) and the National Institute on Minority Health and Health Disparities (NIMHD). The authors also wish to thank the staffs and participants of the JHS.

The views expressed in this manuscript are those of the authors and do not necessarily represent the views of the National Heart, Lung, and Blood Institute; the National Institutes of Health; or the U.S. Department of Health and Human Services.

### CFS

The Cleveland Family Study (CFS) was designed to examine the genetic basis of sleep apnea in 2,534 African-American and European-American individuals from 356 families. Index probands with confirmed sleep apnea were recruited from sleep centers in northern Ohio, supplemented with additional family members and neighborhood control families [{Redline1995}]. Four visits occurred between 1990 and 2006; in the first 3, data were collected in participants’ homes while the last occurred in a clinical research center (2000 - 2006). Measurements included sleep apnea monitoring, blood pressure, anthropometry, spirometry and other related phenotypes. Blood samples (overnight fasting, before bed and following an oral glucose tolerance test), nasal and oral ultrasound, and ECG were also obtained during the 4th exam. Institutional Review Board approval and signed informed consent was obtained for all participants.

#### Ethics statement

Cleveland Family Study was approved by the Institutional Review Board (IRB) of Case Western Reserve University and Mass General Brigham (formerly Partners HealthCare). Written informed consent was obtained from all participants.

#### Acknowledgements

The Cleveland Family Study has been supported in part by National Institutes of Health grants [R01-HL046380, KL2-RR024990, R35-HL135818, and R01-HL113338].

## Description of phenotypes used in the analysis

**Table S1:** Phenotypes used in the analyses and their codes. Phenotypes harmonized by TOPMed DCC used in this study either as target phenotypes, covariates, or variables required for removal criteria.

| Phenotype | Transformation if applied | Role | Code in the TOPMed phenotype database |
| --- | --- | --- | --- |
| Sex |  | Adjusting covariate | annotated_sex_1 |
| Age |  | Adjusting covariate | age_at_height_baseline_1 |
| Race / Ethnicity | Only kept samples where the two race phenotypes agree | Adjusting covariate or stratification | race_us_1 |
|  | Only kept samples where the two race phenotypes agree | Adjusting covariate or stratification | hispanic_or_latino_1 |
| Principal Components 1-5 |  | Adjusting covariate | EV1, EV2, EV3, EV4, EV5 |
| Height |  | Phenotype | height_baseline_1 |
| Lipid Lowering medication usage |  | variable | lipid_lowering_medication_1 |
| Total Cholesterol | Log-transformed, only kept samples where no lipid medication was used | Phenotype | total_cholesterol_1 |
| Triglycerides | Log-transformed, only kept samples where no lipid medication was used | Phenotype | triglycerides_1 |
| Blood Pressure medication usage |  | variable | antihypertensive_meds_1 |
| Systolic Blood Pressure | Add +15 to value if blood pressure lowering medications were used | Phenotype | bp_systolic_1 |
| Sleep Duration |  | Phenotype | sleep_duration_1 |

**Table S2:** Summary statistics of phenotypes used in a secondary analysis in which the training dataset included related individuals. Mean, Median and percent of missing data for the phenotypes (Triglycerides, Total Cholesterol, Systolic Blood Pressure, Sleep Duration and Height) and covariates (sex and age) used in this study. All the traits are presented for the whole database as well as broken down by race (Black, White, and Hispanic/Latino). This training set excludes any related individuals above 3rd degree.

|  | Black<br>(N=4264) | Hispanic/Latino<br>(N=5915) | White<br>(N=9878) | Overall<br>(N=20057) |
| --- | --- | --- | --- | --- |
| **Sex** |  |  |  |  |
| Male | 1780 (41.7%) | 2582 (43.7%) | 4535 (45.9%) | 8897 (44.4%) |
| Female | 2484 (58.3%) | 3333 (56.3%) | 5343 (54.1%) | 11160 (55.6%) |
| **Age** |  |  |  |  |
| Mean (SD) | 50.6 (17.9) | 49.0 (13.8) | 54.1 (15.7) | 51.8 (15.8) |
| Median [Min, Max] | 53.0 [9.00, 92.0] | 50.0 [18.0, 86.0] | 55.0 [5.00, 98.0] | 53.0 [5.00, 98.0] |
| **Triglycerides** |  |  |  |  |
| Mean (SD) | 105 (63.2) | 137 (98.1) | 132 (83.3) | 129 (86.3) |
| Median [Min, Max] | 90.0 [16.0, 873] | 115 [20.0, 1670] | 112 [17.0, 1600] | 109 [16.0, 1670] |
| Missing | 1859 (43.6%) | 1060 (17.9%) | 2225 (22.5%) | 5144 (25.6%) |
| **Total Cholesterol** |  |  |  |  |
| Mean (SD) | 202 (41.5) | 201 (43.3) | 209 (39.1) | 205 (41.1) |
| Median [Min, Max] | 198 [81.0, 450] | 198 [62.0, 526] | 207 [77.8, 594] | 203 [62.0, 594] |
| Missing | 1859 (43.6%) | 1060 (17.9%) | 2225 (22.5%) | 5144 (25.6%) |
| **Systolic Blood Pressure** |  |  |  |  |
| Mean (SD) | 125 (20.7) | 121 (17.3) | 118 (17.8) | 120 (18.5) |
| Median [Min, Max] | 121 [73.0, 246] | 119 [77.0, 218] | 115 [67.0, 227] | 118 [67.0, 246] |
| Missing | 1073 (25.2%) | 1321 (22.3%) | 2285 (23.1%) | 4679 (23.3%) |
| **Sleep Duration** |  |  |  |  |
| Mean (SD) | 6.40 (1.44) | 7.71 (1.50) | 6.99 (1.15) | 7.14 (1.45) |
| Median [Min, Max] | 6.00 [1.00, 16.5] | 7.79 [2.00, 13.4] | 7.00 [1.00, 15.0] | 7.00 [1.00, 16.5] |
| Missing | 1196 (28.0%) | 319 (5.4%) | 3990 (40.4%) | 5505 (27.4%) |
| **Height** |  |  |  |  |
| Mean (SD) | 169 (9.47) | 163 (9.25) | 168 (9.75) | 167 (9.89) |
| Median [Min, Max] | 168 [125, 207] | 162 [134, 194] | 168 [109, 203] | 166 [109, 207] |

**Table S3:**
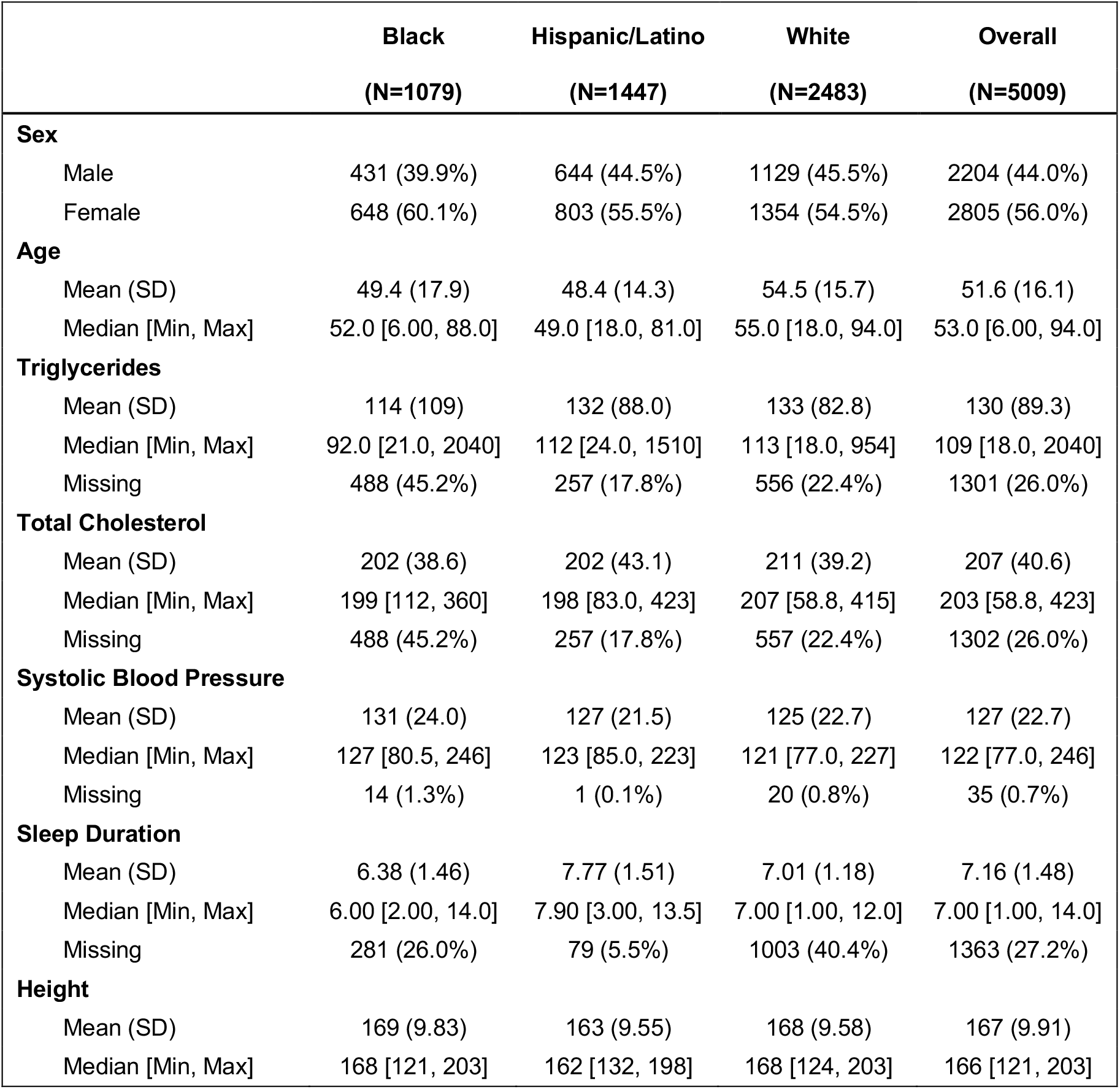
Summary statistics of phenotypes in the testing dataset. Mean, Median and percent of missing data for the phenotypes (Triglycerides, Total Cholesterol, Systolic Blood Pressure, Sleep Duration and Height) and covariates (sex and age) used in this study. All the traits are presented for the whole database as well as broken down by race (Black, White, and Hispanic/Latino). The test set excludes any related individuals above 3rd degree to itself or any of the training datasets.

|  | Black<br>(N=1079) | Hispanic/Latino<br>(N=1447) | White<br>(N=2483) | Overall<br>(N=5009) |
| --- | --- | --- | --- | --- |
| **Sex** |  |  |  |  |
| Male | 431 (39.9%) | 644 (44.5%) | 1129 (45.5%) | 2204 (44.0%) |
| Female | 648 (60.1%) | 803 (55.5%) | 1354 (54.5%) | 2805 (56.0%) |
| **Age** |  |  |  |  |
| Mean (SD) | 49.4 (17.9) | 48.4 (14.3) | 54.5 (15.7) | 51.6 (16.1) |
| Median [Min, Max] | 52.0 [6.00, 88.0] | 49.0 [18.0, 81.0] | 55.0 [18.0, 94.0] | 53.0 [6.00, 94.0] |
| **Triglycerides** |  |  |  |  |
| Mean (SD) | 114 (109) | 132 (88.0) | 133 (82.8) | 130 (89.3) |
| Median [Min, Max] | 92.0 [21.0, 2040] | 112 [24.0, 1510] | 113 [18.0, 954] | 109 [18.0, 2040] |
| Missing | 488 (45.2%) | 257 (17.8%) | 556 (22.4%) | 1301 (26.0%) |
| **Total Cholesterol** |  |  |  |  |
| Mean (SD) | 202 (38.6) | 202 (43.1) | 211 (39.2) | 207 (40.6) |
| Median [Min, Max] | 199 [112, 360] | 198 [83.0, 423] | 207 [58.8, 415] | 203 [58.8, 423] |
| Missing | 488 (45.2%) | 257 (17.8%) | 557 (22.4%) | 1302 (26.0%) |
| **Systolic Blood Pressure** |  |  |  |  |
| Mean (SD) | 131 (24.0) | 127 (21.5) | 125 (22.7) | 127 (22.7) |
| Median [Min, Max] | 127 [80.5, 246] | 123 [85.0, 223] | 121 [77.0, 227] | 122 [77.0, 246] |
| Missing | 14 (1.3%) | 1 (0.1%) | 20 (0.8%) | 35 (0.7%) |
| **Sleep Duration** |  |  |  |  |
| Mean (SD) | 6.38 (1.46) | 7.77 (1.51) | 7.01 (1.18) | 7.16 (1.48) |
| Median [Min, Max] | 6.00 [2.00, 14.0] | 7.90 [3.00, 13.5] | 7.00 [1.00, 12.0] | 7.00 [1.00, 14.0] |
| Missing | 281 (26.0%) | 79 (5.5%) | 1003 (40.4%) | 1363 (27.2%) |
| **Height** |  |  |  |  |
| Mean (SD) | 169 (9.83) | 163 (9.55) | 168 (9.58) | 167 (9.91) |
| Median [Min, Max] | 168 [121, 203] | 162 [132, 198] | 168 [124, 203] | 166 [121, 203] |

## Supplementary results

**Figure S1.**
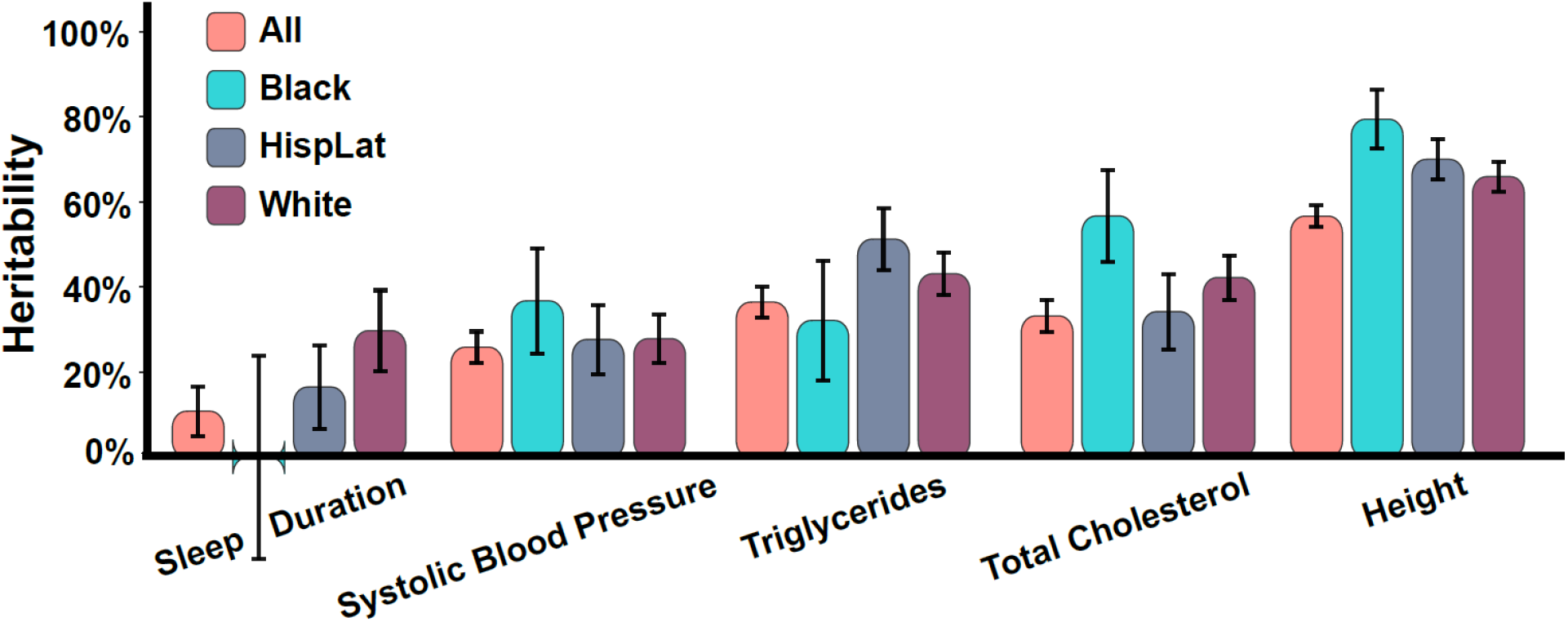
Heritability of Phenotypes by Race/Ethnicity. The heritability of each phenotype for each race/ethnic group.

**Table S4.** Clumping the SNPs prior to performing the XGBoost ensemble model does not significantly change results. The percentage of variance explained in the validation set when the model is trained on the clumped SNPs rather than all SNPs.

| Phenotype | Held-Out Validation PVE for XGBoost Ensemble Model – Clumped SNPs |
| --- | --- |
| Triglycerides | 10.6% |
| Total Cholesterol | 14.6% |
| Systolic Blood Pressure | 2.7% |
| Sleep Duration | 0.46% |
| Height | 21.5% |

**Table S5.**
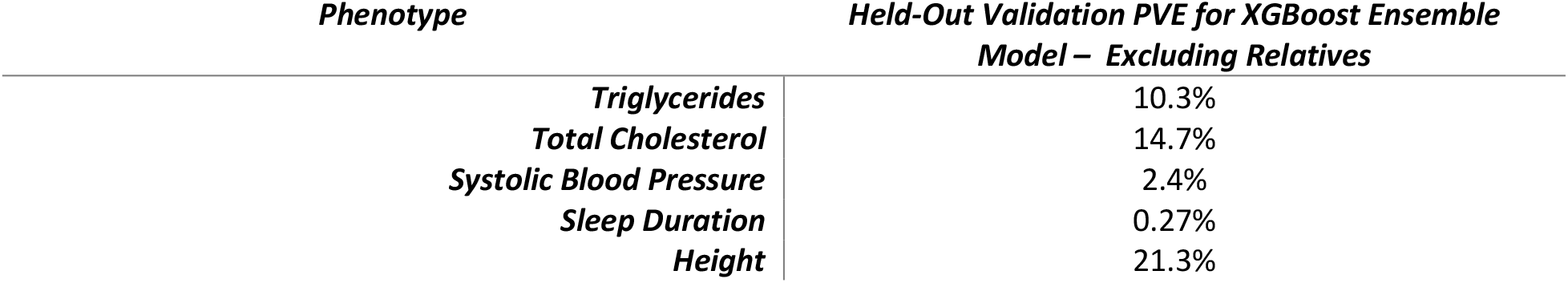
Excluding relatives from the training and validation datasets does not significantly change results. The percentage of variance explained int eh validation set when the model is trained and validated on the subset of subjects that are not related (using the kinship coefficient to define related individuals).

| Phenotype | Held-Out Validation PVE for XGBoost Ensemble Model – Excluding Relatives |
| --- | --- |
| Triglycerides | 10.3% |
| Total Cholesterol | 14.7% |
| Systolic Blood Pressure | 2.4% |
| Sleep Duration | 0.27% |
| Height | 21.3% |

**Table S6.** Results of the PRS model without including the genetic PCs as covariates. The results are slightly lower than the results of the PRS model that does include the genetic PCs as covariates.

| *Phenotype* | *Held-Out Validation PVE for PRS Model – Without*<br>*PCs as Covariates* |
| --- | --- |
| *Triglycerides* | 6.2% |
| *Total Cholesterol* | 8.5% |
| *Systolic Blood Pressure* | 1.2% |
| *Sleep Duration* | 0.06% |
| *Height* | 14.0% |

**Table S7.** Results of the LASSO model when performing cross-validation for the optimal regularization term with respect to the LASSO loss function, rather than the joint-training scheme with XGBoost. Results are slightly higher for the LASSO model, but are not directly comparable to the XGBoost models as they include different variants.

| *Phenotype* | *Held-Out Validation PVE for LASSO Model – Cross-*<br>*Validation for Optimal Regularization Term* |
| --- | --- |
| *Triglycerides* | 6.8% |
| *Total Cholesterol* | 10.6% |
| *Systolic Blood Pressure* | 1.6% |
| *Sleep Duration* | 0.027% |
| *Height* | 10.5% |

**Table S8.** Selected parameters in cross-validation in the main multi-ethnic analysis. Regularization parameters and number of SNPs selected through cross-validation. α refers to the regularization parameter in the LASSO. θ refers to the number of gradient boosted trees.

| *Phenotype* | *XGBoost Alone (and*<br>*LASSO with XGBoost*<br>*variants)* |  |  | *XGBoost with PRS* |  |  | *LASSO with*<br>*Cross-Validation* |  | *PRS* |  |
| --- | --- | --- | --- | --- | --- | --- | --- | --- | --- | --- |
| | $\alpha$ | SNPs | $\theta$ | $\alpha$ | SNPs | $\theta$ | $\alpha$ | SNPs | P-Value | SNPs |
| *Triglycerides* | 0.007 | 432 | 872 | 0.008 | 304 | 1000 | 0.018 | 14 | 0.001 | 7500 |
| *Total Cholesterol* | 0.003 | 455 | 842 | 0.003 | 455 | 1554 | 0.002 | 1162 | 0.001 | 6960 |
| *Systolic Blood Pressure* | 0.258 | 72 | 244 | 0.17 | 346 | 544 | 0.146 | 85 | 0.001 | 8066 |
| *Sleep Duration* | 0.021 | 91 | 373 | 0.013 | 666 | 385 | 0.008 | 1708 | 0.001 | 4876 |
| *Height* | 0.057 | 2193 | 985 | 0.083 | 916 | 995 | 0.017 | 14163 | 0.001 | 22461 |

**Table S9.** Number of SNPs selected through cross-validation in the race/ethnic-specific XGBoost models. Number of SNPs selected through cross-validation in the Black, Hispanic/Latino, and White XGBoost models that included the C+T PRS.

| *Phenotype* | *Black* | *Hispanic/Latino* | *White* |
| --- | --- | --- | --- |
| *Sleep Duration* | 271 | 309 | 341 |
| *Systolic Blood Pressure* | 533 | 516 | 307 |
| *Triglycerides* | 743 | 98 | 685 |
| *Total Cholesterol* | 833 | 442 | 617 |
| *Height* | 959 | 2433 | 725 |

**Table S10.** Support for Figure 2. Non-linear model consistently outperforms linear ones for prediction of multiple complex phenotypes in human cohort.

| *Phenotype* | *PRS* | *LASSO* | *XGBoost Alone* | *XGBoost with PRS* | *Heritability* |
| --- | --- | --- | --- | --- | --- |
| *Sleep Duration* | 0.4% | 0.0% | 0.4% | 0.5% | 10.8% |
| *Systolic Blood Pressure* | 1.8% | 0.0% | 1.5% | 2.6% | 25.9% |
| *Triglycerides* | 6.4% | 5.9% | 7.8% | 10.5% | 36.4% |
| *Total Cholesterol* | 8.1% | 10.1% | 12.3% | 15.0% | 33.2% |
| *Height* | 17.2% | 6.5% | 7.2% | 21.7% | 56.6% |

**Table S11.** Support for Figure 3. Model performance depends on the population with XGBoost consistently outperforming PRS.

| *Phenotype* | *PRS* |  |  | *XGBoost with PRS* |  |  |
| --- | --- | --- | --- | --- | --- | --- |
|  | *White* | *Black* | *Hispanic / Latino* | *White* | *Black* | *Hispanic / Latino* |
| *Sleep Duration* | 0.7% | 0.1% | 0.2% | 0.3% | 0.0% | 0.2% |
| *Systolic Blood Pressure* | 1.8% | 1.2% | 2.6% | 2.5% | 1.8% | 3.8% |
| *Triglycerides* | 5.6% | 6.7% | 8.4% | 8.7% | 5.5% | 14.0% |
| *Total Cholesterol* | 7.4% | 7.0% | 9.6% | 12.9% | 8.1% | 17.4% |
| *Height* | 25.8% | 8.1% | 17.0% | 28.1% | 8.1% | 22.9% |

**Table S12.**
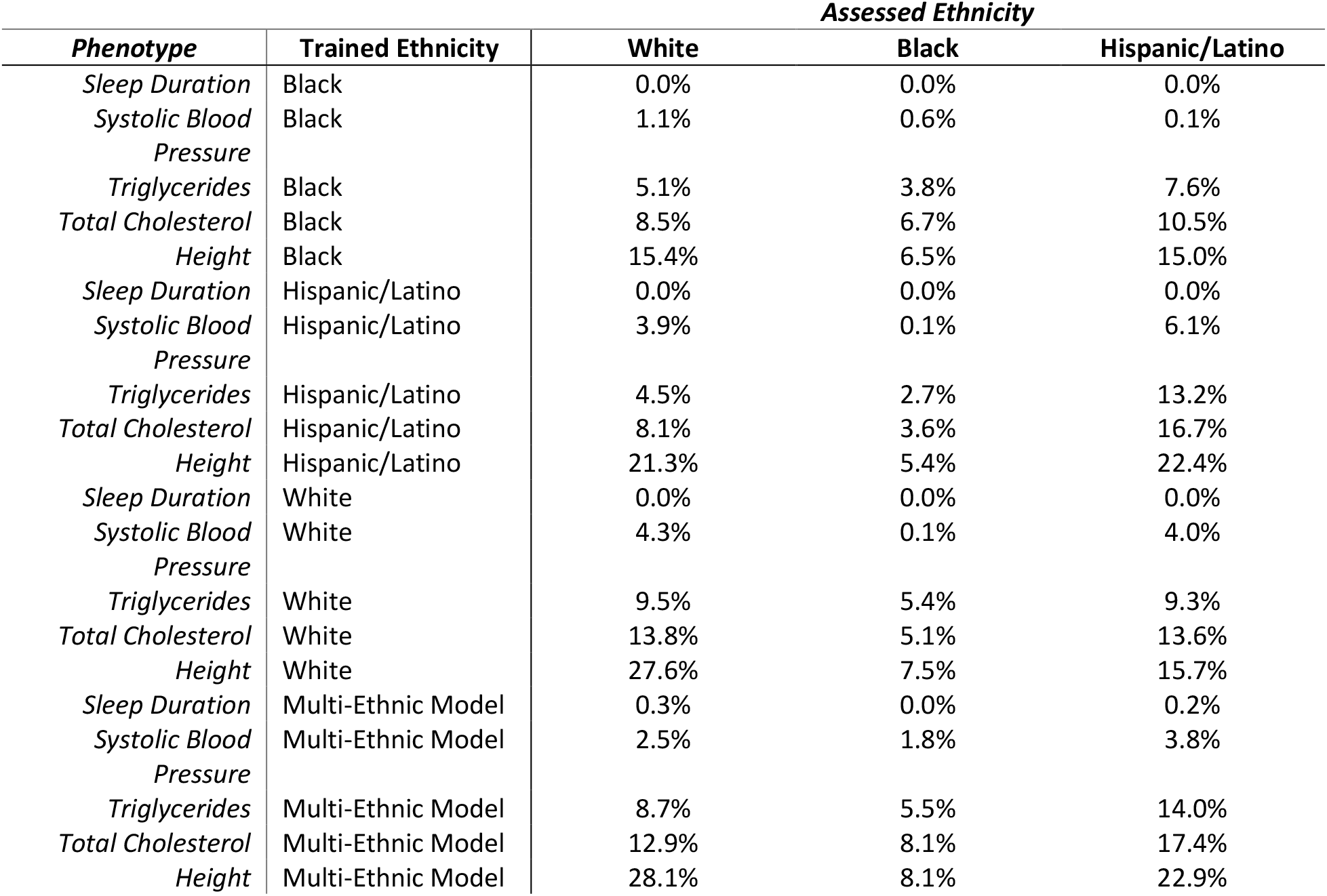
Support for Figure 4. Multi-ethnic XGBoost model performs on par with the race/ethnic-specific models.

